# Malaria species prevalence among asymptomatic individuals in four regions of Mainland Tanzania

**DOI:** 10.1101/2023.12.28.23300584

**Authors:** Zachary R. Popkin Hall, Misago D. Seth, Rashid A. Madebe, Rule Budodo, Catherine Bakari, Filbert Francis, Dativa Pereus, David J. Giesbrecht, Celine I. Mandara, Daniel Mbwambo, Sijenunu Aaron, Abdallah Lusasi, Samwel Lazaro, Jeffrey A. Bailey, Jonathan J. Juliano, Julie R. Gutman, Deus S. Ishengoma

**Affiliations:** Institute for Global Health and Infectious Diseases, University of North Carolina, Chapel Hill, NC, USA; National Institute for Medical Research, Dar es Salaam, Tanzania; National Institute for Medical Research, Tanga Center, Tanga, Tanzania; The Connecticut Agricultural Experiment Station, New Haven, CT, USA; National Malaria Control Programme, Dodoma, Tanzania; Department of Pathology and Laboratory Medicine, Warren Alpert Medical School, Brown University, RI, USA; Center for Computational Molecular Biology, Brown University, RI, USA; Malaria Branch, Global Health Center, Centers for Disease Control and Prevention, Atlanta, GA, USA; Harvard T. H. Chan School of Public Health, Boston, MA; Faculty of Pharmaceutical Sciences, Monash University, Melbourne, VIC, Australia

**Keywords:** malaria, *Plasmodium malariae*, *Plasmodium ovale*, *Plasmodium vivax*, non-falciparum species, Tanzania, asymptomatic malaria

## Abstract

Recent studies point to the need to incorporate non-falciparum species detection into malaria surveillance activities in sub-Saharan Africa, where 95% of malaria cases occur. Although *Plasmodium falciparum* infection is typically more severe, diagnosis, treatment, and control for *P. malariae*, *P. ovale* spp., and *P. vivax* may be more challenging. The prevalence of these species throughout sub-Saharan Africa is poorly defined. Tanzania has geographically heterogeneous transmission levels but an overall high malaria burden. In order to estimate the prevalence of malaria species in Mainland Tanzania, 1,428 samples were randomly selected from 6,005 asymptomatic isolates collected in cross-sectional community surveys across four regions and analyzed via qPCR to detect each *Plasmodium* species. *P. falciparum* was most prevalent, with *P. malariae* and *P. ovale* spp. detected at lower prevalence (<5%) in all four regions. *P. vivax* was not detected. Malaria elimination efforts in Tanzania will need to account for these non-falciparum species.

---

Tanzania has one of the highest malaria burdens in the world, accounting for 4.1% of global malaria deaths in 2021[1]. While most malaria cases in Tanzania and elsewhere in sub-Saharan Africa are caused by *Plasmodium falciparum*, four other *Plasmodium* species (*P. vivax*, *P. malariae*, *P. ovale curtisi*, and *P. ovale wallikeri*) are present to varying degrees. There is also data to suggest that these species have higher prevalence than previously known, and may become more prevalent as *P. falciparum* is controlled and ultimately eliminated[2–6] in line with the WHO goal of a 90% reduction in global malaria burden by 2030[7]. Non-falciparum malaria may require different control measures, due to major differences in biology, including different anopheline vectors with different seasonal peaks[8], relapse and/or chronic infections[9,10], lower parasitemia[11], and higher rates of asymptomatic infection[8].

Previous work in Mainland Tanzania has characterized non-falciparum prevalence in schoolchildren (5-16 years)[6] and non-falciparum positivity rates among symptomatic patients[12]. In schoolchildren, *P. ovale* spp. prevalence (24%) was similar to that of *P. falciparum* (22%)[6], while in symptomatic patients, *P. falciparum* was much more abundant than non-falciparum malaria, although *P. ovale* spp. positivity rates surpassed 5% in seven of ten regions[12]. In both studies, *P. malariae* was less common than either *P. falciparum* or *P. ovale* spp., and *P. vivax* was rare[6,12]. In this study, we characterize the prevalence of all malaria species among asymptomatic individuals across all ages in three regions with moderate and high malaria transmission intensity, and in children under five in one region with high transmission.

The study protocol was approved by the Tanzanian Medical Research Coordinating Committee (MRCC) of the National Institute for Medical Research (NIMR) and involved approved standard procedures for informed consent and sample deidentification. Additional details are described elsewhere[13]. Deidentified samples were considered non-human subjects’ research at the University of North Carolina and Brown University.

A random subset of 1,428 dried blood spot (DBS) samples were drawn from a total of 5,860 asymptomatic samples. 694 samples were drawn from a total of 2,647 collected from all age groups during cross-sectional community surveys in Kigoma (n=252/878, high transmission), Ruvuma (n=186/741, high transmission), and Tanga (n=256/1,028, moderate transmission) regions during the Molecular Surveillance of Malaria in Tanzania (MSMT) project in 2021[13]. The random subset was representative of the regional sample distribution (*X*^2^ (3,2) = 2.43, p (2 df) = 0.3, **Table S1**), but not representative of the age group distribution (*X*^2^ (3,2) = 10.46, p (2 df) = 0.005, **Table S2**), or the sex distribution (*X*^2^ (2,2) = 43.65, p (1 df) < 0.001, **Table S3**). An additional 734 samples were drawn from 3,213 collected from children under five during cross-sectional household surveys for the group antenatal care project (GANC)[14,15] in Geita in 2021.

The molecular analyses used to detect *Plasmodium* spp. in each sample are described in detail elsewhere[12]. Briefly, we performed a separate 18S qPCR assay for each species, which allows for both the detection of each species as well as a semi-quantitative parasitemia estimate. For each region, we calculated prevalence for each species, including both single-species and mixed-species infections. Regional-level maps of prevalence for each species were created using the R package *sf* (version 1.0-9) based on shape files available from GADM.org and naturalearthdata.com accessed via the R package *rnaturalearth* (version 0.3.2)[16].

Variation in species-specific prevalence by region and age group (young children <5 years, school-aged children 5-16 years, and adults >16 years, as previously described[12]) was assessed for significance with generalized linear models or ANOVA, as appropriate, in R. Excluding the Geita participants who were all under five and whose ages were not recorded, the median age of the remaining 694 participants was 20 years (IQR 8-47) with a range of 6 months to 87 years. Including the Geita participants, children (≤16 years old) constituted 74.2% of participants (n=1,060), while adults (>16 years old) constituted the remaining 25.8% (n=368). Young children (<5 years old) comprised 77.9% (n=826) of the child participants, while the remaining 22.1% (n=234) were school-aged (5-16 years old). Sex identifications were available for 694 participants and female-skewed, with 505 female (72.8%) and 189 male participants (27.2%). 21.1% (n=301) of sampled individuals were RDT-positive.

Among all 1,428 samples analyzed, *P. falciparum* was detected in 34.2% (n=488, 95% CI: 31.7%-36.7%), *P. malariae* in 1.5% (n=22, 95% CI: 0.99%-2.4%), and *P. ovale* spp. in 3.4% (n=49, 95% CI: 2.6%-4.5%). *P. vivax* was not detected. *P. malariae* infections were nearly evenly split between single-species infections (45.5%, n=10/22) and mixed-species infections with *P. falciparum* (40.9%, n=9/22), with the remaining three (13.6%) being triple infections with *P. falciparum* and *P. ovale* spp. (**Table 1**). In contrast, most *P. ovale* spp. infections were mixed with *P. falciparum* (65.3%, n=32/49), with single-species infections being less common (28.6%, n=14/49) and triple infections comprising the remainder (**Table 1**). *P. malariae* had the highest median parasitemia at 164,080 p/µL (IQR 9,942-1,333,100 p/µL), followed by *P. falciparum* at 55,200 p/µL (2,910-775,000 p/µL) and *P. ovale* spp. at 11,868 p/µL (1,271-70,840 p/µL). However, there was no significant difference in parasitemia by species (ANOVA p=0.9).

**Table 1:** Infection composition proportions for isolates infected with *P. malariae* and *P. ovale* spp.

| <u>Infection Type</u> | <u>Count</u> | <u>Proportion</u> |
| --- | --- | --- |
| *Pm* | 10 | 45.5% |
| *Pm/Pf* | 9 | 40.9% |
| *Pm/Pf/Po* | 3 | 13.6% |
| All *Pm* Infections | 22 |  |
| *Po* | 14 | 28.6% |
| *Po/Pf* | 32 | 65.3% |
| *Pm/Pf/Po* | 3 | 6.1% |
| All *Po* Infections | 49 |  |

\*Sum of proportions may not equal 100% due to rounding.

All three species were detected in each region (**Figure 1**). Geita and Tanga had the highest *P. falciparum* prevalence (37.8% and 36.7%, n=278/734 and 94/256 respectively, **Table S4**). *P. malariae* was relatively rare in all four regions, with the highest prevalence in Tanga (3.1%, n=8/256) and the lowest in Geita (0.7%, n=5/734, **Table S4**). *P. ovale* spp. prevalence was slightly higher than *P. malariae* (3.2%-4.0%, n=6/186–10/252) in all regions except Tanga (2.3%, n=6/256, **Table S4**). There was significant variation (by ANOVA) in prevalence between regions for *P. falciparum* (p<0.001) and *P. malariae* (p=0.04) but not for *P. ovale* spp.

**Figure 1:**
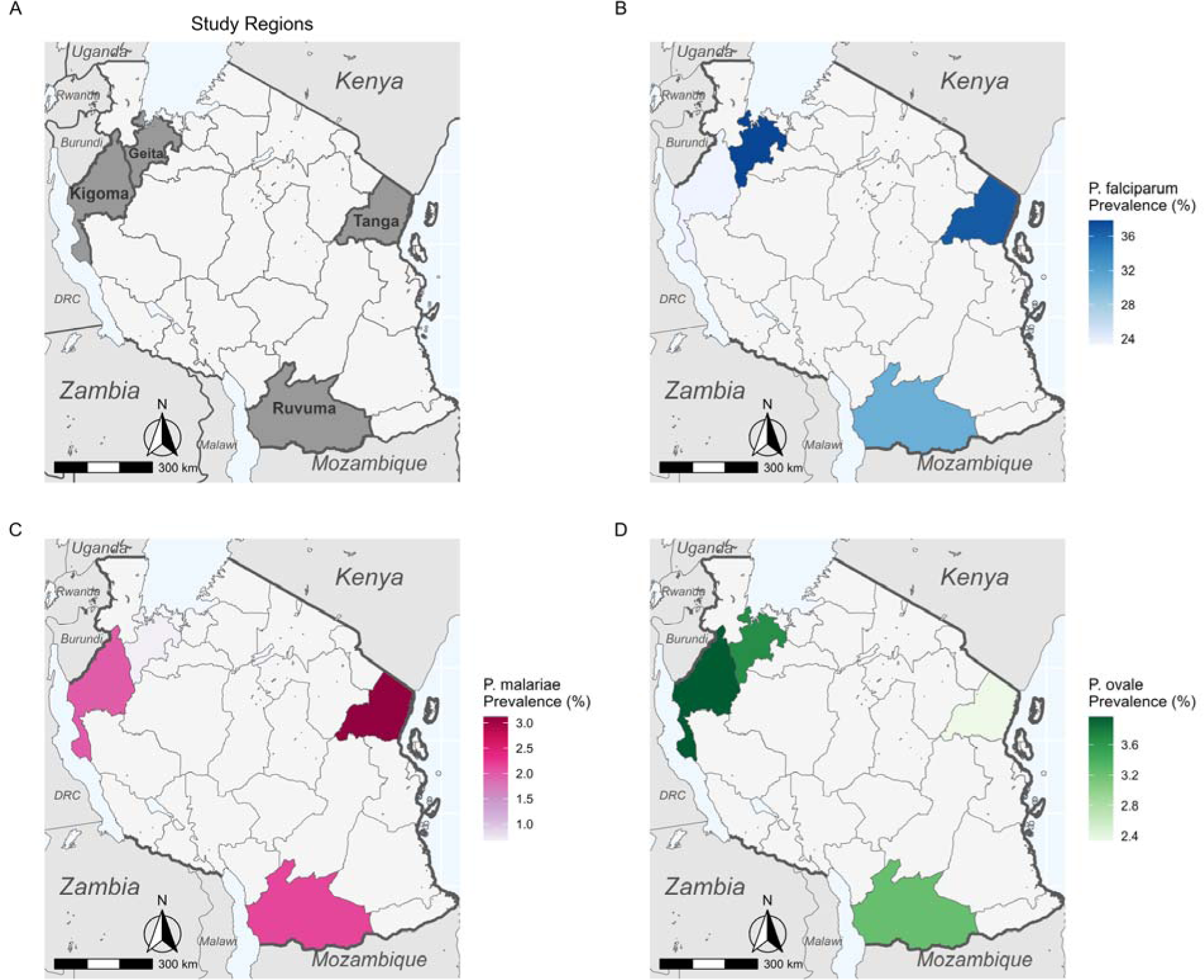
Maps of Tanzania showing A) location of study regions, B) *P. falciparum* regional prevalence, C) *P. malariae* regional prevalence, and D) *P. ovale* spp. regional prevalence. *P. vivax* was not detected, so is not mapped.

While age was a significant (GLM p<0.001) determinant of *P. falciparum* infection, there was no significant effect of age for either *P. malariae* or *P. ovale* spp. Age group was a significant (ANOVA p<0.05) determinant of infection likelihood for both *P. falciparum* and *P. malariae*, but not *P. ovale* spp. (**Figure 2**). While children were significantly (p<0.05) more likely than adults to have *P. falciparum*, adults were more likely to have *P. malariae* (p<0.05, **Figure 2**). There was no significant interaction between age group and region for either *P. falciparum* or *P. malariae*, but the interaction was nearly significant (p=0.055) for *P. ovale* spp.

**Figure 2:**
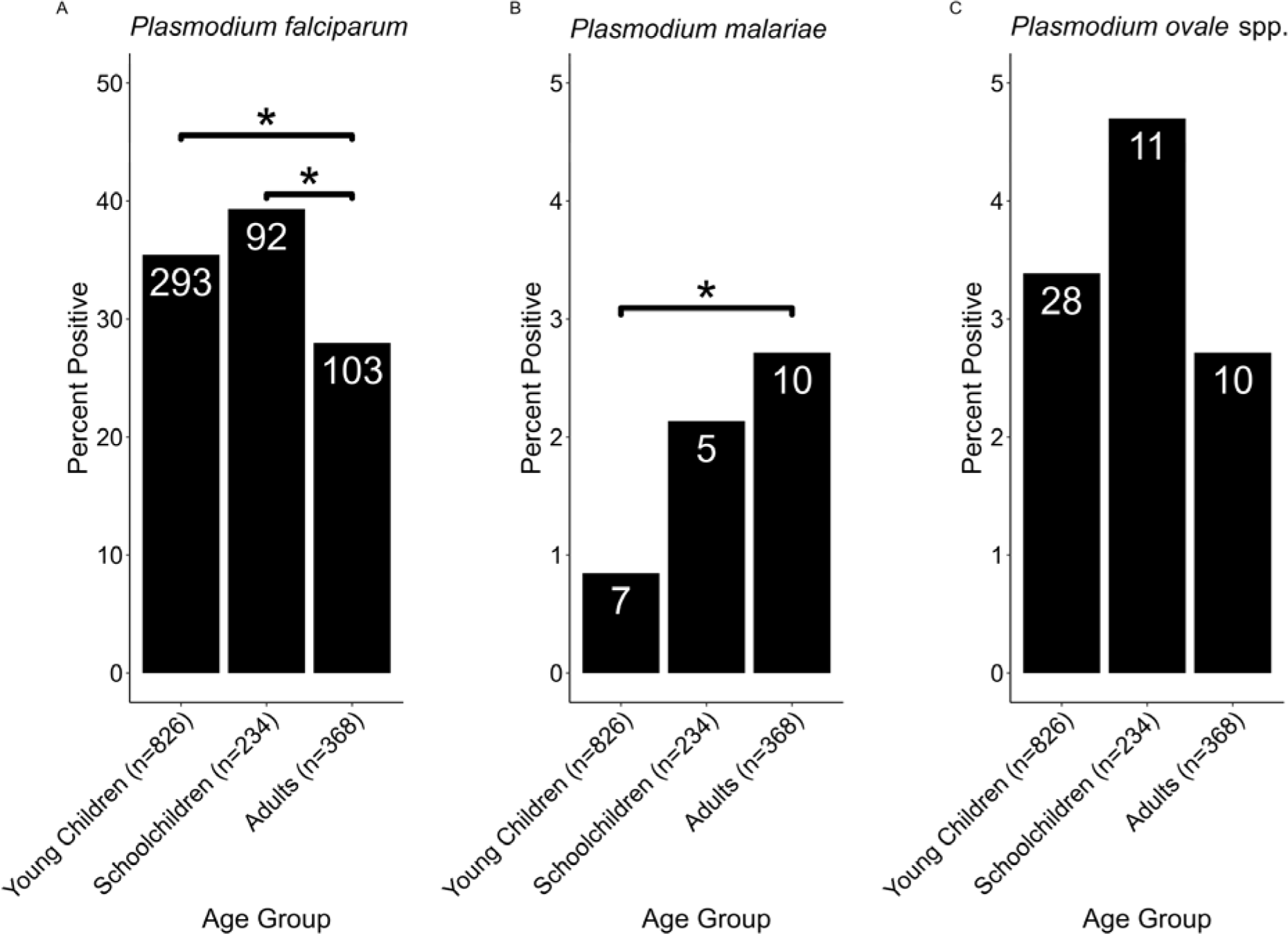
Tukey Analysis of Malaria Species Prevalence by Age Group. A total of 826, 234, and 368 were in the Young Children (<5 years), Schoolchildren (5-16 years), and Adult (>16 years) groups, respectively. The total number of samples per group for each species is shown in the X-axis labels while the number of positive samples for each group is shown in the bar labels. Comparisons marked with a * are significant at the p<0.05 level. **Panel A** shows *P. falciparum* prevalence by age group. Significant pairwise comparisons are marked, while the other is insignificant. **Panel B** shows *P. malariae* prevalence by age group. One significant pairwise comparison is marked, while the others are insignificant. **Panel C** shows *P. ovale* spp. prevalence by age group. No pairwise comparisons are significant.

This study builds on previous research with schoolchildren and clinic patients to describe the prevalence of different malaria species within four regions of Mainland Tanzania. Although *P. falciparum* is the most prevalent species, both *P. malariae* and *P. ovale* spp. prevalence surpasses 3% in at least one region, and could increase as *P. falciparum* is locally eliminated. In contrast to a 2017 study on schoolchildren, which found *P. ovale* spp. prevalence to be similar to *P. falciparum*[6], we found *P. ovale* spp. prevalence to be much lower than that of *P. falciparum*. In addition, the schoolchildren survey mostly identified *P. ovale* spp. as single-species infections and *P. malariae* as mixed with *P. falciparum*[6], whereas we found similar proportions of single-species and mixed-species *P. malariae* infections and most of our *P. ovale* spp. samples were mixed with *P. falciparum*. However, our sample sizes are small and may not necessarily be representative of the full picture, particularly in Geita where samples were only collected from children under 5 years old. In addition, our study only includes four regions, only one of which (Tanga) overlaps with the previous study, and we include a wider age range.

In Tanga, we may have found lower *P. ovale* spp. prevalence than the previous study due to the inclusion of adults, who are less likely to test positive for this species, whereas schoolchildren are a major asymptomatic infectious reservoir[17–19], although we did not replicate a significant difference in this study. In addition, the disruptions to malaria control caused by the COVID-19 pandemic, particularly in 2020-2021[20], could have an increase in *P. falciparum* prevalence. Indeed, there was no difference in the malaria prevalence in the villages of Magoda, Mamboleo, and Mpapayu between 2019 (24.9%) and 2021 (24.5%, unpublished data). However, the prevalence dropped to 6.4% in 2022 (unpublished data), so further longitudinal study may clarify the impact of resumed intense *P. falciparum* control. However, the malaria prevalence in Tanga dropped from 34.8% to 26.2% between 2020 and 2021 (unpublished data), so *P. falciparum* control in this region was likely effective during the course of this study.

Unlike in our study of symptomatic patients[12], we did not find schoolchildren to be significantly more likely to test positive for either *P. malariae* or *P. ovale* spp. Although not significant, this trend is present in *P. ovale* spp. prevalences in this study, so the lack of significance in these species is likely an artifact of small sample sizes (**Table 1**, **Figure 2**, **Figure S1**). Our finding that *P. malariae* was significantly more prevalent among adults likely reflects the presence of chronic infections[10].

Although *P. falciparum* remains the most prevalent species in these four regions, *P. malariae* and *P. ovale* spp. are present in all four, whereas *P. vivax* is not detected. Achieving malaria elimination in Tanzania will require ongoing surveillance of these species. While standard treatments successfully clear *P. falciparum*, the 3.4% of patients in this study with *P. ovale* spp. may relapse. This study serves as a complement to previous studies focusing on schoolchildren and symptomatic patients to paint a full picture of the non-falciparum malaria landscape for communities in Mainland Tanzania. Ongoing analysis of samples collected in 2022 and 2023 will allow us to detect temporal trends in prevalence, and forthcoming genomic analysis of *P. malariae* and *P. ovale* spp. isolates from Tanzania will inform our understanding of population structure and diversity in these species.

## Disclaimer

The findings and conclusions in this report are those of the authors and do not necessarily represent the official position of the U.S. Centers for Disease Control and Prevention.

## Acknowledgements

The authors wish to thank participants and parents/guardians of all children who took part in the surveillance. We acknowledge the contribution of the following project staff and other colleagues who participated in data collection and/or laboratory processing of samples: Raymond Kitengeso, Ezekiel Malecela, Muhidin Kassim, Athanas Mhina, August Nyaki, Juma Tupa, Anangisye Malabeja, Emmanuel Kessy, George Gesase, Tumaini Kamna, Grace Kanyankole, Oswald Osca, Richard Makono, Ildephonce Mathias, Godbless Msaki, Rashid Mtumba, Gasper Lugela, Gineson Nkya, Daniel Chale, Richard Malisa, Sawaya Msangi, Ally Idrisa, Francis Chambo, Kusa Mchaina, Neema Barua, Christian Msokame, Rogers Msangi, Salome Simba, Hatibu Athumani, Mwanaidi Mtui, Rehema Mtibusa, Jumaa Akida, Ambele Yatinga, and Tilaus Gustav. We also acknowledge the finance, administrative and logistic support team at NIMR: Christopher Masaka, Millen Meena, Beatrice Mwampeta, Gracia Sanga, Neema Manumbu, Halfan Mwanga, Arison Ekoni, Twalipo Mponzi, Pendael Nasary, Denis Byakuzana, Alfred Sezary, Emmanuel Mnzava, John Samwel, Daud Mjema, Seth Nguhu, Thomas Semdoe, Sadiki Yusuph, Alex Mwakibinga, Rodrick Ulomi and Andrea Kimboi. We are also grateful to the management of the National Institute for Medical Research, National Malaria Control Program and President’s Office-Regional Administration and Local Government (regional administrative secretaries of the 14 regions, and district officials, staff from all 100 HFs and Community Health Workers from the 4 community cross sectional regions). Technical and logistics support from the Bill and Melinda Gates Foundation team is highly appreciated. The following reagents were obtained through BEI Resources, NIAID, NIH: Diagnostic Plasmid Containing the Small Subunit Ribosomal RNA Gene (18S) from *Plasmodium falciparum*, MRA-177; *Plasmodium vivax*, MRA-178; *Plasmodium malariae*, MRA-179; and *Plasmodium ovale*, MRA-180, contributed by Peter A. Zimmerman. Permission to publish the manuscript was sought and obtained from the Director General of NIMR.

## Authors’ contributions

ZRPH, JAB, JJJ, and DSI conceived the study. ZRPH performed computational and epidemiological analyses and wrote the manuscript. MDS, RAM, RB, CB, and DP collected samples, extracted DNA, and performed qPCR analysis. CIM, JAB, JJJ, and DSI oversaw the project. FF and DJG contributed data analysis. JRG oversaw data collection in Geita and assisted with statistical analysis. DM, SA, AL, and SL contributed data from NMCP and facilitated data collection. JAB, JJJ, JRG, and DSI edited the manuscript. All authors read, contributed to, and approved the final manuscript.

## Data availability

Data is available upon reasonable request to the corresponding author.

---

**Figure S1:**
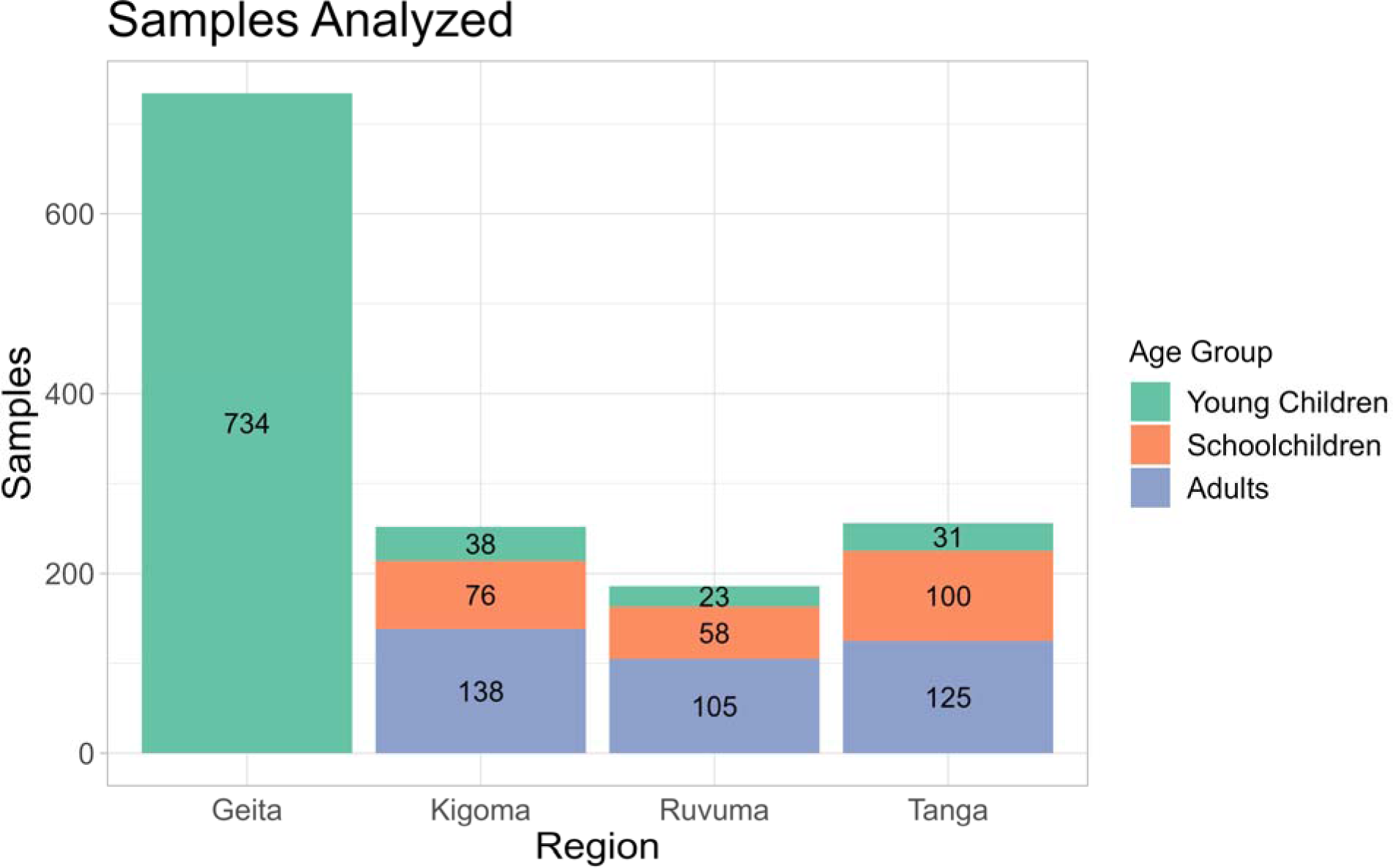
Samples included in analysis by age group and region.

**Supplemental Table 1:** Regional Distribution of Samples in Random Subset and Full Dataset.

| **Region** | **Subset** | **Full Dataset** |
| --- | --- | --- |
| Kigoma | 252 (36.3%) | 878 (33.2%) |
| Ruvuma | 186 (26.8%) | 741 (28.0%) |
| Tanga | 256 (36.9%) | 1,028 (38.8%) |
| **Total Samples** | **694** | **2,647** |

**Supplemental Table 2:**
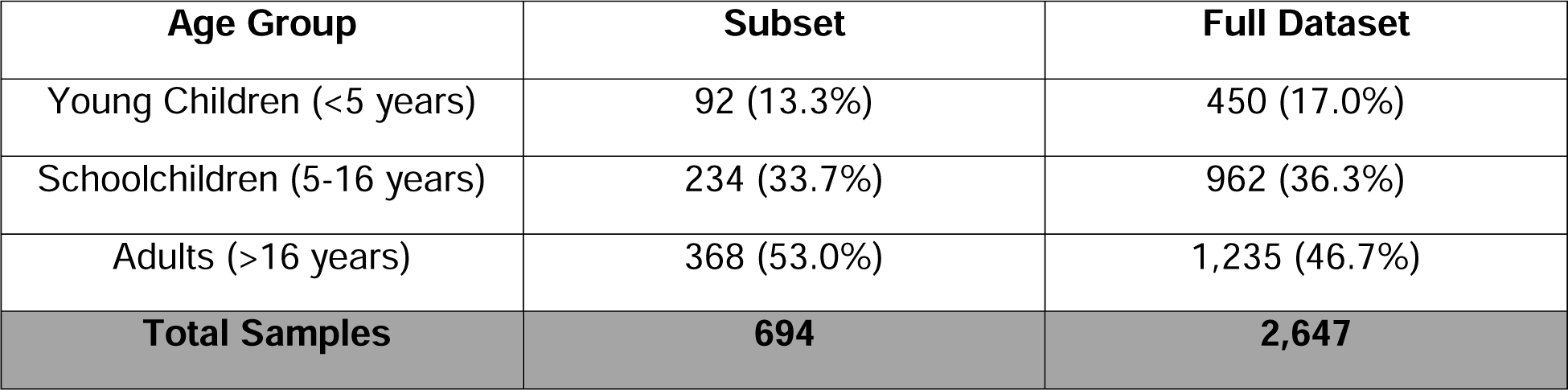
Age Group Distribution of Samples in Random Subset and Full Dataset.

**Supplemental Table 3:** Sex Distribution of Samples in Random Subset and Full Dataset.

| **Sex** | **Subset** | **Full Dataset** |
| --- | --- | --- |
| Female | 505 (72.8%) | 1,568 (59.3%) |
| Male | 189 (27.2%) | 1,078 (40.7%) |
| **Total Samples** | **694** | **2,646\*** |

\*Sex was not reported for all individuals.

**Supplemental Table 4:** Species Positivity by Region and Age Group.

| Region | Age Group | Pf-Positive Samples | Pm-Positive Samples | Po-Positive Samples | Total Samples |
| --- | --- | --- | --- | --- | --- |
| Geita* | Young Children | 278 (37.9%) | 5 (0.7%) | 27 (3.7%) | 734 |
|  | Schoolchildren | * | * | * | * |
|  | Adults | * | * | * | * |
|  | **All Ages** | **278 (37.9%)** | **5 (0.7%)** | **27 (3.7%)** | **734** |
| Kigoma | Young Children | 7 (18.4%) | 1 (2.6%) | 1 (2.6%) | 38 |
|  | Schoolchildren | 26 (34.2%) | 2 (2.6%) | 7 (9.2%) | 76 |
|  | Adults | 26 (18.8%) | 2 (1.4%) | 2 (1.4%) | 138 |
|  | **All Ages** | **59 (23.4%)** | **5 (2.0%)** | **10 (4.0%)** | **252** |
| Ruvuma | Young Children | 0 | 0 | 0 | 23 |
|  | Schoolchildren | 22 (37.9%) | 2 (3.4%) | 3 (5.1%) | 58 |
|  | Adults | 35 (33.3%) | 2 (1.9%) | 3 (2.9%) | 105 |
|  | **All Ages** | **57 (30.6%)** | **4 (2.2%)** | **6 (3.2%)** | **186** |
| Tanga | Young Children | 8 (25.8%) | 1 (3.2%) | 0 | 31 |
|  | Schoolchildren | 44 (44%) | 1 (1%) | 1 (1%) | 100 |
|  | Adults | 42 (33.6%) | 6 (4.8%) | 5 (4%) | 125 |
|  | **All Ages** | **94 (36.7%)** | **8 (3.1%)** | **6 (2.3%)** | **256** |

\*Only children under five were enrolled in the study in Geita. Schoolchildren are defined as ages
5-16, and adults are all those >16 years.

